# Association of overweight/obesity and insulin resistance with activation of circulating innate lymphoid cells in women after gestational diabetes mellitus

**DOI:** 10.1101/2025.01.15.25320580

**Authors:** Julia Sbierski-Kind, Stephan Schlickeiser, Lorenzo Semeia, Saori Harada, Eleni Pappa, Javier Villamizar Cujar, Minh-Thuy Katschke, Christina Gar, Andreas Lechner, Andreas L. Birkenfeld, Uta Ferrari, Jochen Seissler

## Abstract

**Introduction:** Women with a history of gestational diabetes mellitus (GDM) are at high risk of developing prediabetes or type 2 diabetes later in life. Recent studies have highlighted the regulation and function of innate lymphoid cells (ILCs) in metabolic homeostasis. However, the multifactorial impact of both overweight/obesity and GDM on the immunological profile of circulating ILCs and the progression to prediabetes are not yet fully elucidated.

**Methods:** Blood samples from 42 women with a history of insulin-treated GDM (GDMi), 33 women with a history of GDM without insulin treatment during pregnancy (GDM), and 45 women after a normoglycemic pregnancy (Ctrl) participating in the ongoing observational PPSDiab study were analyzed by flow cytometry for markers of ILC subsets at the baseline visit (3-16 months postpartum; Visit 1) and 5 years postpartum (58-66 months postpartum; Visit 3).

**Results:** During the first 5 years postpartum, 18 women of the GDMi group (42.8%), 10 women of the GDM group (30.3%), and 8 participants of the Ctrl group (17.8%) developed prediabetes, respectively. Total circulating type 1 innate lymphoid cells (ILC1s) and NK cell numbers as well as percent HLA-DR^+^ ILC1s were increased in GDMi versus GDM and Ctrl women both at the baseline visit and the 5-year follow-up. Although ILC subsets at Visit 1 could not predict the progression from GDM to prediabetes, ILC2 frequency was associated with insulin sensitivity index (ISI), whereas percent HLA-DR^+^ ILC1s were inversely correlated. Moreover, circulating leukocytes and total NK cells were associated with waist circumference and fat mass both at Visit 1 and Visit 3.

**Discussion:** Our findings introduce human ILCs as a potential therapeutic target deserving further exploration.

**Trial registration:** Study ID 300-11.

## 1 Introduction

Gestational diabetes mellitus (GDM), a common metabolic disorder during pregnancy, is characterized by glucose intolerance that typically manifests in the second or third trimester. Affecting 7-10% of pregnancies worldwide (1), it poses significant health risks to both the mother and the developing fetus, including cardiovascular diseases and adverse perinatal outcomes such as preeclampsia, macrosomia, neonatal hypoglycemia, and an increased risk for cesarean delivery (2–5). GDM is also a risk factor of type 2 diabetes mellitus later in life for both the mother and the offspring (6), underscoring the importance of understanding the underlying pathophysiological mechanisms. Previous German studies reported an incidence of diabetes mellitus after GDM, ranging from 5.5% within the first year to 52.7% within 20 years postpartum (7–10). Since 2011, GDM screening (a 50g glucose challenge followed by a 75g oral glucose tolerance test (OGTT) applying the International Association of the Diabetes and Pregnancy Study Groups (IADPSG)) is offered to all pregnant women as part of prenatal care in Germany. The PPSDiab (‘Prediction, Prevention, and subclassification of Type 2 Diabetes’) Study, a prospective, postpartum cohort study, was then started to gain more insight into prediabetes and type 2 diabetes after GDM (11). We recently reported an incidence of 6% for type 2 diabetes and 1% for type 1 diabetes during the first 5 years postpartum, whereas 55% of the study participants cumulatively developed prediabetes (12), indicating that GDM is a strong risk factor for impaired postpartum glucose metabolism.

While, in recent years, the focus has been on metabolic and hormonal changes during pregnancy, emerging research highlights the complex interplay between immune system alterations and the development of GDM and subsequent metabolic disturbances (13–15). Insulin resistance is associated with low-grade inflammation of hypertrophic adipose tissue and promotes type 2 diabetes when exhausted beta cells fail to maintain glucose homeostasis (16,17). Among the emerging areas of interest is the role of innate lymphoid cells (ILCs) in regulating immune responses during pregnancy and their potential impact on the onset and progression of GDM (18). ILCs are critical regulators of tissue homeostasis, inflammation, and metabolic processes (19–22). These cells are classified into three groups (ILC1s, ILC2s, and ILC3s) based on associated cytokine profiles (type 1 - IFNγ; type 2 - IL-4, IL-5, IL-9, IL-13; type 3/17 - IL-17A/F, IL-22) and transcriptional regulators (23). ILCs coordinate various physiological processes, including anti-microbial immune responses, modulating adaptive immunity as well as organ development, homeostasis, and repair (23). Recent work suggests that fully differentiated ILC subsets, including NK cells, derive from CD117^+^ innate lymphoid cell precursors (ILCps) which are mainly found in the circulation but also in tissues (24).

Recent studies have highlighted their involvement in metabolic disorders, including type 1 and type 2 diabetes. In particular, group 2 innate lymphoid cells (ILC2s) are present in metabolic tissues in both humans and mice and promote browning and other adipose tissue remodeling (25,26). As integral regulators of adipose tissue type 2 immunity, they promote eosinophils and alternatively activated macrophages as well as regulatory T cells (27) to maintain an anti-inflammatory environment. Impaired ILC2 response results in glucose intolerance in mice, whereas restoration of ILC2s in adipose tissues promotes whole-body sensitivity (28,29). Importantly, it has been shown that IFNy-producing group 1 innate lymphoid cells (ILC1s) drive pro-inflammatory macrophage polarization in the adipose tissue and contribute to insulin resistance during diet-induced obesity (30). However, the functional role of ILC subsets in GDM remains largely unexplored. Given the similarities between the pathophysiological mechanisms in GDM and other metabolic disorders, it is probable that ILCs contribute to the development of GDM and subsequent glucose impairment. By elucidating the potential of ILCs to regulate insulin resistance and glucose metabolism in women with a history of GDM, our work aims to deepen our understanding of immune-related mechanisms underlying this condition and potentially identify new biomarkers and treatment strategies.

In this ongoing, observational PPSDiab study, we used multicolor flow cytometry from (1) women after a normoglycemic pregnancy (n=45, ‘Ctrl’); (2) women with a history of GDM and without insulin treatment during pregnancy (n=33, ‘GDM’); and (3) women with a history of GDM and with insulin treatment during pregnancy (n=42, ‘GDMi’) to identify specific immunological alterations, including ILCs, in GDM at the baseline visit after delivery and the 5-year follow-up. We found persistent ILC alterations in GDMi women until the 5-year follow-up and ILC subpopulations were associated with metabolic measures. As such, our data represent an important step towards a better understanding of immune-metabolism interactions and will open novel avenues to investigate human peripheral blood ILC subsets in health and disease.

## 2 Methods and materials

### 2.1 Study design and population

Individuals included in the present analysis were participants in the prospective, monocentric observational cohort PPSDiab study (31). Women with GDM during their last pregnancy with insulin treatment (GDMi group) or without insulin treatment (GDM group) and women after a normoglycemic pregnancy (Ctrl group) were recruited within 3 to 16 months after delivery in the ratio 2:1 from the diabetes center and the obstetrics department of the University Hospital in Munich (LMU Klinikum), Germany between November 2011 and May 2016. The diagnosis of GDM was based on a 75 g oral glucose tolerance test (OGTT) after the 23rd week of gestation. The cut-off values for GDM diagnosis were ≥92 mg/dl (≥5.1mmol/l) after fasting, ≥180 mg/dl (≥10.0 mmol/l) after 1 hour, and ≥153 mg/dl (≥8.5 mmol/l) after 2 hours, according to the recommendations of the IADPSG (32). Normoglycemia during pregnancy was confirmed by an OGTT after the 23rd week of gestation with 75g of glucose (cut-off values for GDM according to IADPSG) or with a 50 g of glucose screening test (cut-off value after 1h < 135mg/dl (<7.5mmol/l) to confirm normoglycemia). The study protocol, inclusion and exclusion criteria have been previously described in detail (33), including alcohol or substance abuse and chronic diseases requiring medication, except for hypothyroidism and mild hypertension. None of the women included in this study experienced subsequent pregnancies between Visit 1 and Visit 3. For the present analysis, peripheral blood mononuclear cells (PBMCs) from the baseline visit (3-16 months after delivery; Visit 1) and the 5-year follow-up (58-66 months postpartum; Visit 3) were available from 120 women, 45 Ctrl participants, 33 GDM patients, and 42 GDMi patients. None of these individuals were treated with antidiabetic drugs after pregnancy. For the GDM group and the GDMi group, annual in-person visits, including OGTTs, were scheduled after the baseline visit. The study participation was terminated for women who developed diabetes mellitus but is still ongoing for the remaining participants.

### 2.2 Anthropometrics and clinical measurements

Height and waist circumference were measured to the nearest 1 cm. Body composition was measured by a bioelectrical impedance analysis scale (Tanita BC-418; Tanita Corporation, Tokyo, Japan). Plasma glucose was measured by a hexokinase method (Glucose HK Gen.3; Roche Diagnostics, Mannheim, Germany), serum insulin was measured by a chemiluminescent immunoassay (DiaSorin LIASON Systems, Saluggia, Italy), and plasma leptin was measured by an enzyme-linked Immunosorbent Assay (ELISA “Dual Range”, Merck Millipore, Darmstadt, Germany). All blood samples, except for post-load glucose and insulin during OGTT, were determined after an overnight fast (12 hours). Blood lipids (enzymatic caloric test, Roche Diagnostics, Mannheim, Germany) and TSH were quantified in a central laboratory (34). Low-density lipoprotein (LDL) cholesterol was calculated by the Friedewald equation. Plasma glycated hemoglobin (HbA1c) was measured by high-performance liquid chromatography (VARIANT II TURBO HbA1c Kit, Bio-Rad Laboratories, Hercules, California, USA). Resistin was measured by an ELISA (Quantikine ELISA, R&D Systems, Wiesbaden-Nordenstadt, Germany), and Adiponectin was measured by a radioimmunoassay (RIA, Merck Millipore, Darmstadt, Germany).

### 2.3 OGTT

The 75 g OGTT was conducted with a five-point measurement of plasma glucose and serum insulin as previously described (35). We used ADA criteria (fasting glucose ≥ 100mg/dl (≥ 5.6mmol/l) and/or 2-hour post-load glucose ≥ 140mg/dl (≥7.8mmol/l)) for the diagnosis of impaired fasting glucose (IFG) and impaired glucose tolerance (IGT). From the OGTT, the insulin sensitivity index (ISI) was calculated according to Matsuda and DeFronzo (36). The Homeostatic Model Assessment for Insulin Resistance (HOMA-IR) was calculated as fasting glucose (mmol/l) x fasting insulin (mmol/l) /22.5.

### 2.4 Blood samples

Peripheral blood samples from all participants were collected in potassium-EDTA-coated blood collection tubes (S-Monovette 7.5ml of K3 EDTA; Sarstedt) and were immediately processed at University Hospital, LMU, Munich, Germany. For isolation of PBMCs, 7.5ml of fresh EDTA blood were collected and centrifuged by density gradient within 30h using the Hypaque-Ficoll method, as previously described (31). Isolated cells were counted and mixed with 2 ml of dimethyl sulfoxide (DMSO; Carl Roth GmbH + Co. KG) containing 10% of fetal calf serum (FCS, FBS Superior; Biochrom, Cambourne). PBMCs were stored in liquid nitrogen for cryopreservation before further analyses.

### 2.5 Flow cytometry

For flow cytometry, cryopreserved PBMCs were thawed in a 37°C water bath, pipetted into Iscove’s Modified Dulbecco’s Medium (IMDM) supplemented with 10% FCS medium, and washed several times by centrifugation, as previously described (37). Three to six million cells per sample were stained with antibodies to surface antigens **(Supplementary Table 2)** for 30 minutes at 4°C, followed by washing with FACS buffer (PBS containing 3% FCS and 0.05% NaN3). The cells were fixed with 2% paraformaldehyde for 10 minutes, washed again with FACS buffer, and resuspended in FACS buffer. Samples were immediately acquired on a BD LSR Fortessa X-20 flow cytometer. Fluorochrome compensation was performed with single-stained UltraComp eBeads (Invitrogen, Life Technologies Corp., Carlsbad, CA). Samples were FSC-A/SSC-A gated to exclude debris, followed by FSC-H/FSC-A gating to select single cells and Zombie NIR fixable or DAPI to exclude dead cells. Innate lymphoid cells were identified as lineage negative (CD1a^-^, CD14^-^, CD19^-^, CD34^-^, CD94^-^, CD123^-^, FcER1a^-^, TCRab^-^, TCRgd^-^, BDCA2^-^), CD45^+^, CD161^+^, CD127^+^, as indicated. The full gating strategy is shown in Fig. S1 and was adapted from previous work (38). Analysis of flow cytometry data was performed using FlowJo version 10.7 software (TreeStar, Ashland, OR, USA) and compiled using Prism (GraphPad Software, La Jolla, CA).

### 2.6 Unsupervised data analysis

Cytobank (39) was used for initial manual gating of Lineage-negative cells and ILC subsets, using the same gating strategy as described above. Lineage-negative cells were subjected to dimensionality reduction using Cytobank opt-SNE with default hyperparameters and following embedding markers with normalized scales Cytobank arcsinh transformation: CD117, CD127, CD161, CD45RA, CD56, CRTH2, HLA-DR, and SLAMF1, as described previously(37). To perform downstream statistical analyses in R (http://www.r-project.org/) and visualize t-SNE maps, events within ILC subsets were exported from Cytobank as tab-separated values containing compensated and transformed marker expression levels as well as t-SNE coordinates and metacluster assignment. T-SNE plots were generated after subsampling each sample to contain a maximum of 2500 events. High-resolution group differences were visualized by calculating Cohen’s D for a given comparison across the t-SNE map. Adaptive 2D histograms were generated by using the probability binning algorithm available through the R *flowFP* package (40). A single binning model was created on collapsed data from all samples, by recursively splitting the events at the median values along the two t-SNE dimensions. After applying the arcsine-square-root transformation for proportions, the group-difference effect sizes were calculated for each bin using the cohen.d function of the *effsize* package. Analyses were performed using R version 4.1.1, available free online at https://www.r-project.org.

### 2.7 Statistical analysis

All metric and normally distributed variables are reported as mean ± standard deviation (SD) unless otherwise noted. A two-sided *p* value of <0.05 was considered statistically significant. Data distributions were tested for normality using the Shapiro–Wilk test. For comparisons between three groups, one-way analysis of variance (ANOVA) with Tuke’s multiple comparisons test was used for normally distributed variables and Kruskal-Wallis test with Dunn’s multiple comparisons test was used for non-normally distributed variables. For comparisons between two groups, two-sided Student’s t-test was used for normally distributed variables and Mann-Whitney U test was used for non-normally distributed variables.

Correlation analyses were performed using Spearman or Pearson correlation coefficients (*p*). Each symbol reflects one individual. We also performed linear regression analyses with HOMA-IR (Table 2) and logistic regression analyses with impaired glucose metabolism at the 5-year follow-up visit (IFG and/or IGT; Table 3) as the dependent variable and insulin sensitivity (ISI; Table 3), HOMA-IR, and ILC subsets (Table 2 and 3) at the first postpartum visit as independent variables adjusted for age and BMI. To address non-normality of the distribution, all variables were log-transformed for regression analyses. In a further, exploratory step, we evaluated whether ISI was correlated to any of the ILC parameters, adjusting for age and BMI.

The regression models considered ISI as dependent variable, ILC parameters as independent variable, and age and BMI as covariates. For all regression analyses, we tested for multicollinearity between independent variables. Data were analyzed using Matlab R2019b, SPSS (IBM Corp. Released 2021. IBM SPSS Statistics for Windows, Version 28.0. Armonk, NY: IBM Corp) or Prism version 8 (GraphPad Software, La Jolla, CA).

## 3 Results

### 3.1 Baseline and follow-up characteristics of study participants

A total of 120 participants of the ongoing observational PPSDiab study were included in the final analysis. Peripheral blood mononuclear cells (PBMCs) from the baseline visit (Visit 1; 3 to 16 months after delivery) and the 5-year follow-up (Visit 3; 58-66 months after delivery) were available from 45 control participants after a normoglycemic pregnancy (Ctrl group), 33 patients with a history of GDM and without insulin treatment during pregnancy (GDM group), and 42 patients with a history of GDM with insulin treatment during pregnancy (GDMi group). Circulating ILC subsets were analyzed and investigated for their correlations with metabolic measures, as shown in the study outline in **Figure 1A**. The baseline characteristics of the study subjects are shown in **Table 1**; follow-up characteristics at Visit 3 are shown in **Supplementary Table 1**. Briefly, the mean ± SD age of all participants at baseline was 36 ± 4 years and none of them had pre-existing diabetes. Several metabolic measures, including BMI, waist circumference, fat mass, ISI, and HbA1c were significantly different between Ctrl participants and the GDMi group, whereas GDM women did not differ from the Ctrl group. Triglycerides and Leptin levels in GDMi women were higher compared to both Ctrl and GDM women. Total leukocyte numbers were elevated in GDMi compared to GDM or Ctrl participants, underscoring chronic inflammation as key feature of obesity and insulin resistance (17). Similarly, at Visit 3, BMI, waist circumference, fat mass, HbA1c, Triglycerides, and leukocyte numbers were all significantly different between GDMi and Ctrl participants, but not GDM women. Only 2 women of the Ctrl group (4.4%) had IFG and/or IGT at Visit 1, whereas 8 (17.8%) of them suffered from either IFG (15.6%) or IGT (2.2%) at Visit 3. Ten out of 33 (30.3%) GDM participants had IFG (15.1%), IGT (9.1%), or both (6.1%) at Visit 1, which did not change at Visit 3. Importantly, 38.1% of GDMi participants were diagnosed with IFG (19.0%), IGT (16.7%), or both (2.4%) 3 to 16 months after delivery, and 42.8% had IFG (21.4%), IGT (9.5%), or both (11.9%) at Visit 3 **(Table 1, Supplementary Table 2 and Figure 1B)**. The prevalence of prediabetes was similar to previous data from the PPSDiab study (12), although none of the participants of the current analysis were diagnosed with type 1 or type 2 diabetes. At Visit 1, HOMA-IR was significantly elevated in GDMi compared to both GDM and Ctrl participants; concomitantly, ISI was decreased in GDMi patients **(Figures 1C and D)**. In summary, GDM patients treated with insulin during pregnancy presented with a worse metabolic profile both at Visit 1 and Visit 3, including significantly higher BMI compared to GDM women without insulin treatment and Ctrl participants, and were at higher risk to develop prediabetes.

**Figure 1.**
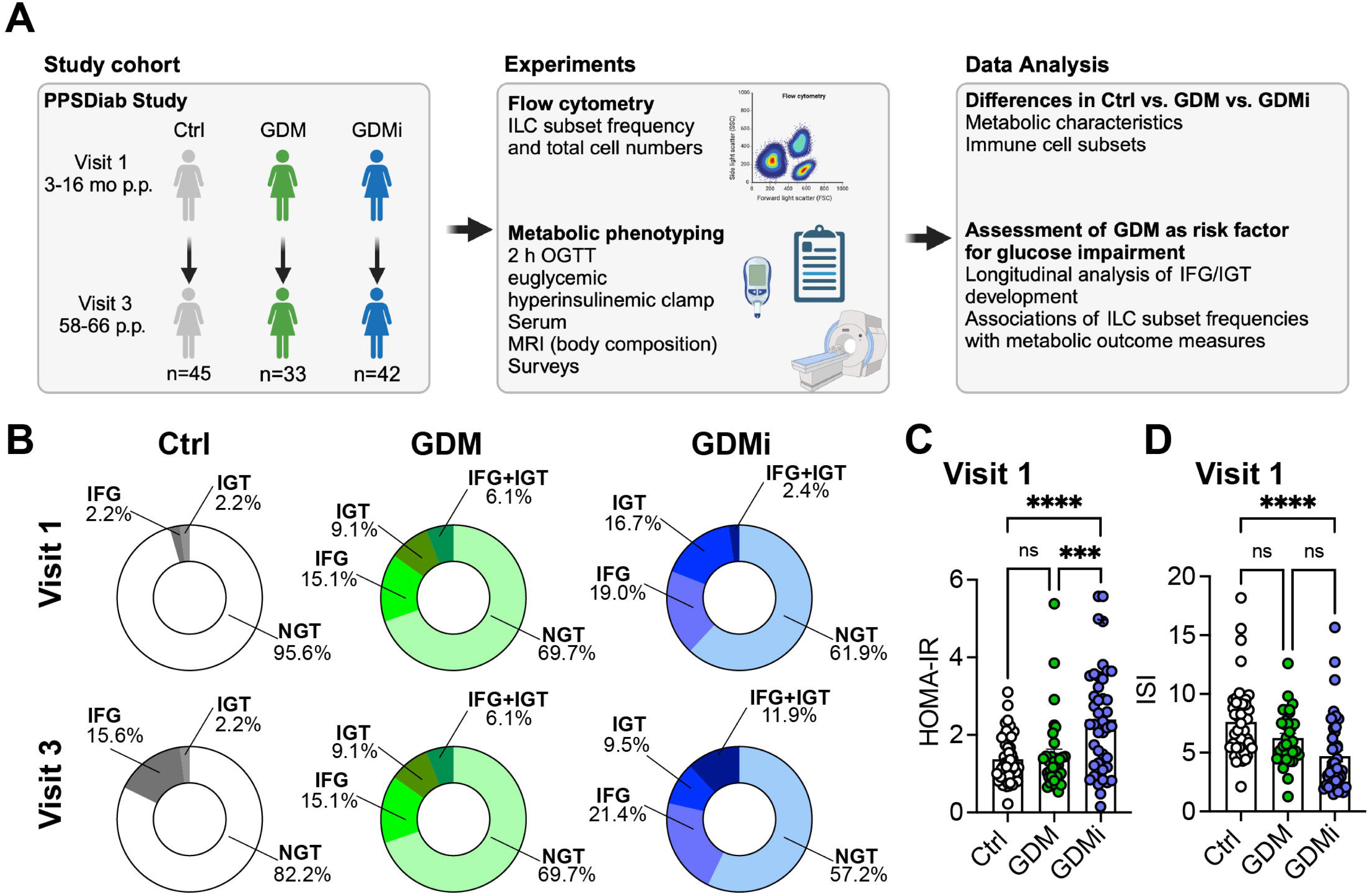
Clinical characteristics of study cohorts. **(A)** Overview of study cohorts and methods. The figure is partly created with BioRender.com. **(B)** Prevalence of impaired fasting glucose (IFG), impaired glucose tolerance (IGT) or normal glucose tolerance (NGT) for healthy control females with normoglycemic pregnancies (Ctrl), females post gestational diabetes mellitus (GDM), and females post gestational diabetes mellitus with insulin treatment during pregnancy (GDMi) at Visit 1 (3-16 months postpartum) and Visit 3 (58-66 months postpartum) displayed as ring charts. Further characteristics are detailed in Table 1. **(C, D)** Homeostatic Model Assessment for Insulin Resistance (HOMA-IR) (C) and insulin sensitivity index (ISI) (D) of Ctrl, GDM or GDMi study participants at Visit 1. Postpartum (p.p.). Bar graphs indicate mean (±SE), n=33-45 individuals per group, one-way ANOVA with Tukey post-hoc test (c, d); ***p ≤ 0.001, ****p ≤ 0.0001.

**Table 1.** Baseline characteristics of study participants.

| Group | Control | GDM | GDMi | Adjusted p-value (Ctrl. vs. GDM) | Adjusted p-value (Ctrl. vs. GDMi) | Adjusted p-value (GDM vs. GDMi) |
| --- | --- | --- | --- | --- | --- | --- |
| No. of subjects | 45 | 33 | 42 |  |  |  |
| Age (years) | 35.58 ± 4.19 | 35.58 ± 3.84 | 36.67 ± 4.14 | >0.999 | 0.430 | 0.486 |
| BMI (kg/m<sup>2</sup>) | 23.46 ± 3.83 | 22.48 ± 3.08 | 26.35 ± 5.98 | 0.616 | 0.010 | 0.001 |
| Waist circumference (cm) | 77.4 ± 8.90 | 75.33 ± 5.83 | 83.69 ± 11.49 | 0.593 | 0.005 | 0.0005 |
| Fat mass (kg) | 20.78 ± 7.64 | 18.24 ± 5.93 | 25.77 ± 10.80 | 0.395 | 0.019 | 0.0007 |
| ISI | 7.62 ± 3.17 | 6.24 ± 2.27 | 4.71 ± 3.16 | 0.106 | <0.001 | 0.0715 |
| HbA1c (%) | 5.16 ± 0.32 | 5.21 ± 0.32 | 5.41 ± 0.28 | 0.743 | 0.0008 | 0.0193 |
| HDL cholesterol (mg/dl) | 65.04 ± 13.20 | 64.61 ± 11.40 | 63.02 ± 17.36 | 0.990 | 0.789 | 0.8841 |
| LDL cholesterol (mg/dl) | 105.69 ± 31.41 | 100.06 ± 25.52 | 106.24 ± 31.30 | 0.690 | 0.996 | 0.6484 |
| Triglycerides (mg/dl) | 66.36 ± 16.74 | 67.00 ± 29.96 | 91.38 ± 60.34 | 0.997 | 0.013 | 0.028 |
| Leptin (ng/ml) | 9.19 ± 6.89 | 8.62 ± 5.32 | 14.40 ± 8.49 | 0.937 | 0.003 | 0.002 |
| Adiponectin (ng/ml) | 12.90 ± 4.95 | 14.83 ± 7.63 | 11.55 ± 5.82 | 0.356 | 0.553 | 0.057 |
| TSH (uIU/ml) | 2.21 ± 1.44 | 2.4 ± 1.97 | 1.83 ± 0.99 | 0.837 | 0.464 | 0.227 |
| Resistin (ng/ml) | 6.72 ± 2.67 | 7.05 ± 3.52 | 7.04 ± 3.01 | 0.887 | 0.881 | 0.999 |
| Leukocytes (10<sup>9</sup>/L) | 5.17 ± 1.16 | 5.38 ± 1.25 | 6.33 ± 1.51 | 0.760 | <0.001 | 0.007 |
| Glucose metabolism (%) |  |  |  |  |  |  |
| NGT | 43 (95.6) | 23 (69.7) | 26 (61.9) |  |  |  |
| IFG | 1 (2.2) | 5 (15.1) | 8 (19.0) |  |  |  |
| IGT | 1 (2.2) | 3 (9.1) | 7 (16.7) |  |  |  |
| IFG+IGT | 0 (0) | 2 (6.1) | 1 (2.4) |  |  |  |
| T2DM | 0 (0) | 0 (0) | 0 (0) |  |  |  |

Data are presented as means (± standard deviation). Adjusted p-values were obtained from post-hoc tests adjusted for multiple comparisons following a one-way analysis of variance (ANOVA) or Kruskal-Wallis test. Visit 1 (3-16 months postpartum) and Visit 3 (58-66 months postpartum). GDM, gestational diabetes mellitus. BMI, body mass index. IFG, impaired fasting glucose. IGT, impaired glucose tolerance. NGT, normal glucose tolerance. ISI, insulin sensitivity index.

### 3.2 Multidimensional flow cytometry characterization of ILCs in GDM

To investigate peripheral blood ILC levels via flow cytometry in GDM compared to normoglycemic pregnancies, we used a well-established gating strategy (38) **(Supplementary Figure 1)**. Lin^-^CD127^+^ ILCs were further defined as CD117^-^CRTH2^-^ ILC1, CRTH2^+^ ILC2s, and CD117^+^ ILCps. CD45RA^+^ ILCs were recently described as quiescent, naïve-like cell subsets, lacking proliferative activity (41). We used CD56 as a marker of activated or ILC3/NK cell-committed ILCps and HLA-DR as activation marker. We performed stochastic neighbor embedding analysis to visualize multiple dimensions in simple two-dimensional plots and to compare flow cytometry data between groups **(Figures 2A and B)**. As described before, CRTH2 was mainly expressed on ILC2s, whereas CD117 was expressed on ILCps **(Figure 2A)**. Interestingly, HLA-DR was highly expressed on distinct subsets of ILC1s and ILC2s with increased expression in GDMi patients compared to Ctrl participants **(Figures 2A and B)**. Furthermore, CD45RA^dim^ ILC1s were decreased in the GDMi group compared to healthy controls and an ILC1 subset with low CD45RA expression was higher in GDM women compared to the Ctrl group **(Figure 2B)**. Next, we evaluated total numbers and frequencies of circulating ILC subsets and NK cells in women with GDM with or without insulin treatment during pregnancy compared to normoglycemic pregnancies.

**Figure 2.**
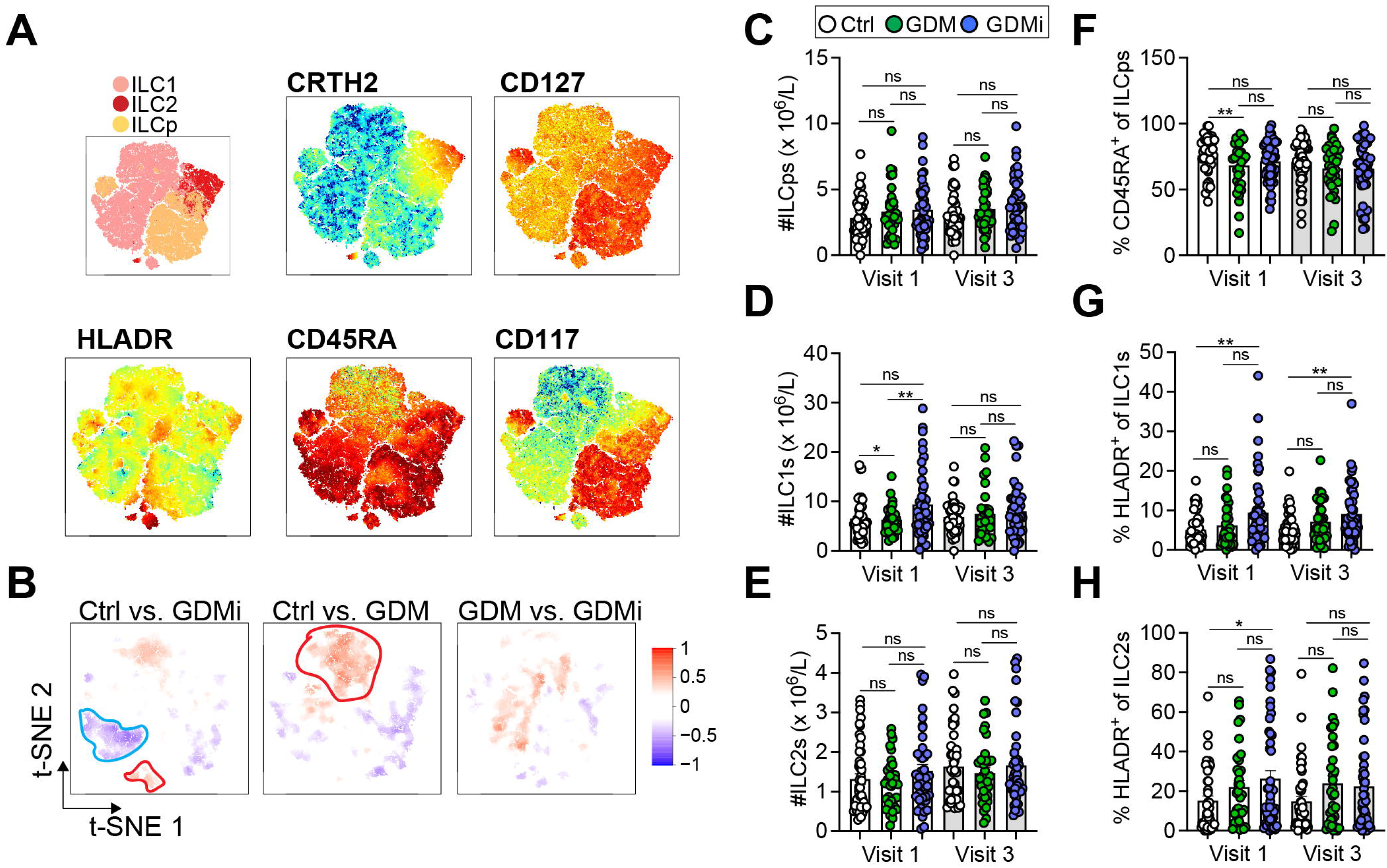
**(A)** Relative expression intensities of surface markers in peripheral blood mononuclear cells (PBMCs) of healthy control females with normoglycemic pregnancies (Ctrl), females post gestational diabetes mellitus (GDM), and females post gestational diabetes mellitus with insulin treatment during pregnancy (GDMi) at Visit 1 (3-16 months postpartum) (combined Ctrl, GDM, and GDMi samples) shown as t-SNE analysis. **(B)** High-dimensionality reduction analysis of innate lymphoid cells (ILCs, gated as lymphocytes, singlets, and CD45^+^CD3^-^Lin^-^CD127^+^ cells as shown in Supplementary Fig. 1 from PBMCs) of Ctrl, GDM, and GDMi study participants at Visit 1 (3-16 months postpartum). High-resolution group differences were visualized by calculating Cohen’s D for a given comparison across the t-SNE map. Residual plot showing differences between maps. Phenotypes within red circles indicate higher expression and phenotypes within blue circles indicate lower expression in respective samples. Analysis is based on flow cytometry data from 45 Ctrl, 33 GDM, and 42 GDMi samples. **(C-H)** Quantifications showing total innate lymphoid progenitor cells (ILCps) (C), group 1 innate lymphoid cells (ILC1s) (D), group 2 innate lymphoid cells (ILC2s) (e), percent CD45RA^+^ of ILCps (F), HLADR^+^ of ILC1s (G), and HLADR^+^ of ILC2s (H). Bar graphs indicate mean (±SE), n=33-45 individuals per group, one-way ANOVA with Tukey post-hoc test (C-H); *p ≤ 0.05, **p ≤ 0.01. See also Supplementary Fig. 2.

While we did not observe significant changes in total numbers of ILC2s and ILCps between groups, total NK cells (Visit 1 and Visit 3) and ILC1s (Visit 1) were elevated in GDMi patients as compared to GDM and Ctrl study participants **(Supplementary Figures 2A, Figures 2 C-E)**. Frequencies of ILCps, ILC1s, and ILC2s were not different between groups **(Supplementary Figures 2B-D)**. However, naïve CD45RA^+^ ILCps were lower in GDM patients compared to Ctrl participants at Visit 1 but were not different at Visit 3 **(Figure 2F)**. In parallel with decreased frequencies of CD45RA^+^ ILCps in GDM, we observed an increase in HLA-DR expression in ILC1s and ILC2s of patients with GDM and insulin treatment during pregnancy at Visit 1 and increased frequencies of HLA-DR^+^ ILC1s at Visit 3 **(Figures 2G and H)**. Moreover, frequencies of both NK^bright^ and NK^dim^ cells were elevated in GDMi patients as compared to the Ctrl group, but not in the GDM group, supporting immune cell activation in metabolically unhealthy patients **(Supplementary Figures 2E and I)**. We could not detect significant differences in frequencies of ILC1s, CD45RA^+^ ILC2s and CD45RA^+^ ILC1s, CD117^+^ ILC2s, or CD56^+^ ILCps between GDMi, GDM, and Ctrl participants at both Visit 1 and Visit 3 **(Supplementary Figures 2F, G, J-L)**. However, HLA-DR^+^ ILCps were significantly elevated in GDMi patients at Visit 3, mirroring activation across ILC subsets **(Supplementary Figure 2H)**. These findings suggest that circulating activated ILC1s are involved in insulin resistance, whereas circulating ILC2s might be protective, as previously suggested (26).

### 3.3 Association of metabolic measures with circulating ILC and NK cell frequencies in GDM

Next, we evaluated whether the immune cell alterations detected in patients with a history of GDM and with insulin treatment during pregnancy were related to metabolic measures for insulin sensitivity and body metabolism both within the first year postpartum (Visit 1) and at Visit 3 by using exploratory data analysis. We found that total ILC1 numbers and percent HLA-DR^+^ ILC1s were inversely correlated with ISI, whereas percent ILC2s were positively correlated **(Figures 3A-C)**, supporting previous findings in mice, where ILC2s promote insulin sensitivity (26). Total ILC1 numbers were positively associated with HOMA-IR; however, we did not observe any correlations between other ILC subsets or NK cells with HOMA-IR **(Figure 3D, data not shown).** Conversely, frequencies of ILCps were negatively associated with HbA1c levels but positively correlated with waist circumference **(Figures 3E and H).** In contrast to ILC1s, both total numbers and levels of ILCps were not different between groups **(Figure 2C and Supplementary Figure 2D)**, suggesting that these cells might play a minor role in the pathogenesis of GDM. However, total numbers of NK cells were positively associated with fat mass and waist circumference both at Visit 1 and Visit 3 **(Figures 3F and G, Supplementary Figures 3 B, E)**, and HLA-DR^+^ ILC1s correlated with waist circumference **(Figure 3I, Supplementary Figure 3C)**. Moreover, leukocyte numbers were associated with both fat mass and waist circumference at Visit 3 **(Supplementary Figures 3A, D)**. In linear regression models, HOMA-IR still associated significantly with total ILC1 numbers as well as percent ILC1s (at Visit 1) after adjusting for age and BMI **(Table 2)**. Together, these data indicate that overweight/obesity and insulin resistance are associated with activated HLA-DR^+^ ILC1s and NK cells but inversely related to ILC2s in women with a history of GDM.

**Figure 3.**
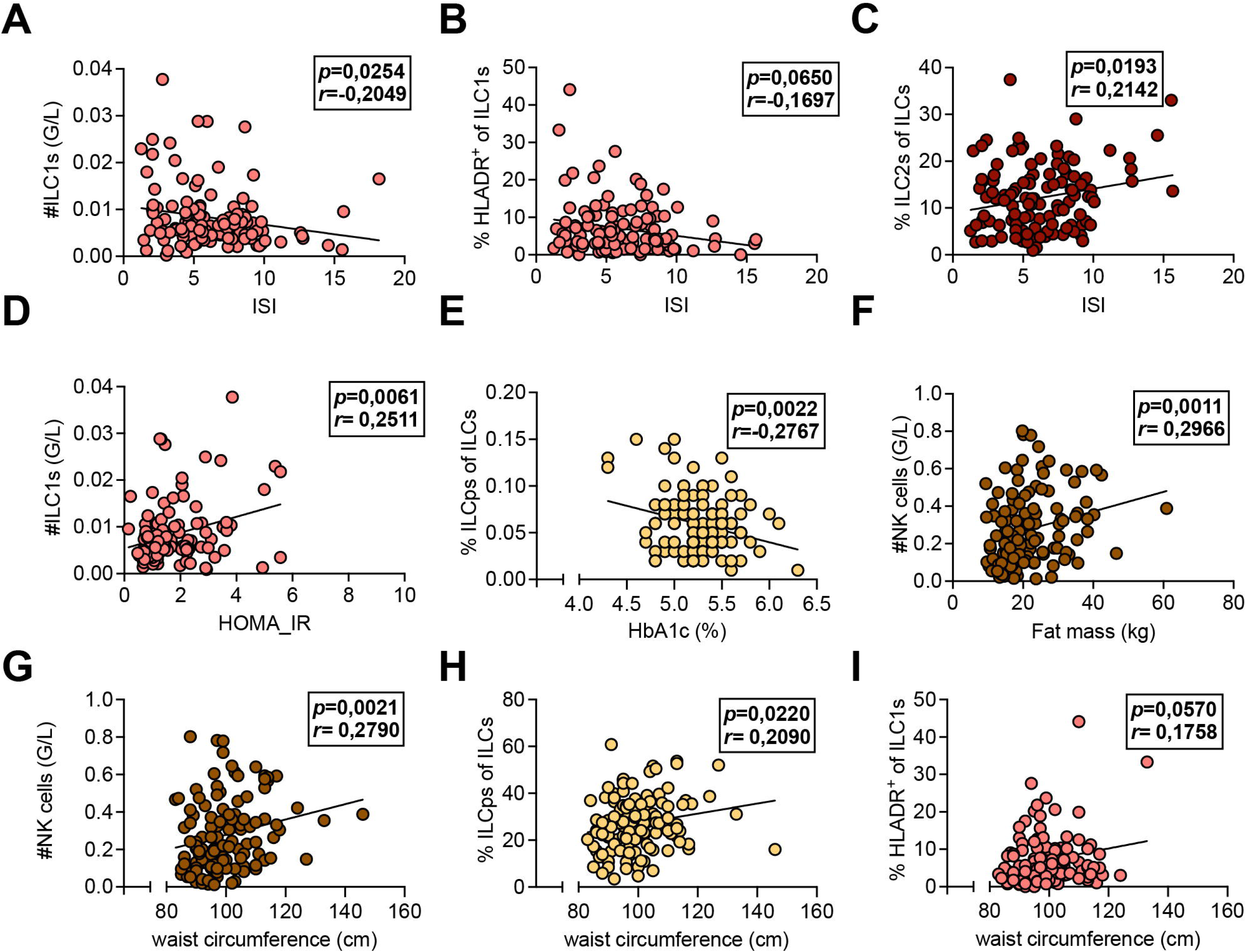
**(A-I)** Associations of systemic leukocyte subpopulations with metabolic measures. Peripheral blood mononuclear cells (PBMCs) isolated from peripheral blood of healthy control females with normoglycemic pregnancies (Ctrl), females post gestational diabetes mellitus (GDM), and females post gestational diabetes mellitus with insulin treatment during pregnancy (GDMi) at Visit 1 (3-16 months postpartum). Subjects were analyzed with multiparameter flow cytometry and the resulting immune cell parameters were correlated with metabolic measures. **(A-C)** Correlations between group 1 innate lymphoid cell (ILC1) counts (A), percent HLADR^+^ of ILC1s (B), and percent group 2 innate lymphoid cells (ILC2s) (C) and the insulin sensitivity index (ISI). **(D)** Correlation between ILC1 counts and Homeostatic Model Assessment for Insulin Resistance (HOMA-IR). **(E)** Correlation between percent innate lymphoid progenitor cells and HbA1c levels. **(F)** Correlation between NK cell counts and fat mass. **(G-I)** Correlations between NK cell counts (G), percent ILCps of ILCs (H), and percent HLADR^+^ of ILC1 (I) with waist circumference. n=33-45 individuals per group. Spearman and Pearson correlation analyses were conducted.

**Table 2.** Linear regression analyses with (log-transformed) Homeostatic Model Assessment for Insulin Resistance (HOMA-IR) at Visit 1 as the dependent variable and (log-transformed) total numbers of frequencies of peripheral blood innate lymphoid cell subsets (log-transformed) as independent variables. Linear regression analysis not adjusted (model 1) and adjusted for age and BMI (model 2).

| ILC subgroups | model 1<sup>a</sup> |  |  | model 2<sup>b</sup> |  |  |
| --- | --- | --- | --- | --- | --- | --- |
| | $\beta$ | R<sup>2</sup> | *p* | $\beta$ | R<sup>2</sup> | *p* |
| #logILCs (G/L) | 8.844 | 0.0243 | 0.0551 | 6.259 | 0.3706 | 0.0917 |
| #logLC1s (G/L) | 11.489 | 0.0343 | 0.0282 | 9.158 | 0.3817 | 0.0294 |
| %logLC1s of CD45<sup>+</sup> cells | 59.356 | 0.0176 | 0.0865 | 63.521 | 0.3846 | 0.0218 |
| %logLC1s of ILCs | 0.2569 | 0.0026 | 0.4007 | 0.4617 | 0.3746 | 0.0605 |

Standardised regression coefficients are shown.
<sup>a</sup>model 1: not adjusted; <sup>b</sup>model 2: adjusted for age and BMI

### 3.4 Association of 5-year follow-up glucose impairment with metabolic measures, circulating ILC and NK cell frequencies in GDM

Next, we asked whether metabolic measures or immune cell parameters (at Visit 1) could predict the risk to develop glucose impairment (IFG and/or IGT) at Visit 3. Using logistic regression models, we found that both HOMA-IR and ISI measured within the first year postpartum (Visit 1) associated significantly with having IFG, IGT, or both at the 5-year follow-up (Visit 3) **(Table 3)**, while circulating ILC subsets or NK cells could not predict glucose impairment (data not shown). Specifically, neither total numbers of ILC1s nor percent ILC1s associated significantly with glucose impairment at Visit 3 **(Table 3)**. Together, we conclude that parameters for evaluating insulin resistance taken in the postpartum period after GDM could help stratifying women at high-risk for the development of glucose impairment and type 2 diabetes.

**Table 3.** Logistic regression analyses with glucose impairment (either impaired fasting glucose (IFG), impaired glucose tolerance (IGT), or both measured during a 75g oral glucose tolerance test (OGTT)) at Visit 3 as the dependent variable and Homeostatic Model Assessment for Insulin Resistance (HOMA-IR), insulin sensitivity index (ISI), total numbers of frequencies of peripheral blood group 1 innate lymphoid cell (ILC1s) subsets (all log-transformed) as independent variables. Logistic regression analysis not adjusted (model 1) and adjusted for age and body mass index (BMI) (model 2).

| **Table 3** Associations between metabolic measures, ILC subgroups and glucose impairment (Visit 3) |  |  |  |  |  |  |
| --- | --- | --- | --- | --- | --- | --- |
| metabolic measures | **model 1<sup>a</sup>** |  |  | **model 2<sup>b</sup>** |  |  |
|  | OR | R2 | *p* | OR | R2 | *p* |
| logHOMA-IR | 6.4738 | 0.0852 | 0.0028 | 6.1387 | 0.0751 | 0.0213 |
| logISI | 0.1628 | 0.1133 | 0.0009 | 0.1591 | 0.15044 | 0.0058 |
| #logLC1s (G/L) | 0.9889 | -0.0081 | 0.7429 | 0.9837 | 0.0248 | 0.6293 |
| %logLC1s of ILCs | 0.9985 | -0.0022 | 0.4155 | 0.9986 | 0.0297 | 0.4668 |

OR, odds ratios for glucose impairment (Visit 3) based on standardised regression coefficients are shown.
<sup>a</sup>model 1: not adjusted; <sup>b</sup>model 2: adjusted for age and BMI

## 4 Discussion

The implications of GDM, the most common medical pregnancy complication worldwide, extend beyond pregnancy, as women diagnosed with GDM are at a higher risk of developing prediabetes and type 2 diabetes later in life (2–5) and their offspring display an increased metabolic risk, probably due to epigenetic reprogramming (42). We previously reported that the cumulative incidence of prediabetes development during the 5-year follow-up after pregnancy was 55% for PPSDiab study participants with a history of GDM, whereas the cumulative incidence of type 2 diabetes was 6% and of type 1 diabetes was 1%, respectively. The prevalence of prediabetes was 37.5% among GDM patients at the 5-year follow-up (12). The lower incidence of both type 2 diabetes and type 1 diabetes compared to previous studies might result from universal screening for GDM, which was established in 2011 in Germany. Consistently, here, we found that 30.3% of women with a history of GDM without insulin treatment during pregnancy and 42.8% of GDM patients with insulin treatment during pregnancy as opposed to 17.8% of women with normoglycemic pregnancies had developed prediabetes at the 5-year follow-up (Visit 3). Importantly, we identified the need for insulin treatment during pregnancies with GDM diagnosis, resulting from the inability to manage blood glucose levels, as risk factor for progression to prediabetes in the postpartum period, thus, suggesting offering specific follow-up care to affected patients.

However, the underlying mechanisms that contribute to this increased risk are multifactorial, involving a complex interplay between metabolic, hormonal, and immunological factors. Although, type 1 inflammation can promote obesity-associated insulin resistance, the precise cellular and temporal contributions, specifically in GDM, are poorly defined. Previous work from the PPSDiab study found no association of circulating CD69^+^ NK cells with overweight/obesity or the metabolic syndrome (31). Similarly, Zhang et al. did not find significant changes in regulatory Treg cell frequencies during the third trimester in GDM patients compared to healthy pregnant women (14). In contrast, several studies reported neutrophil overactivation, increased monocyte activation, elevated B cells and B cell activation, as well as higher numbers of cytotoxic T cells and an elevated Th1 and Th17 response in peripheral blood of GDM patients during pregnancy (15), suggesting that particularly type 1 inflammation might be implicated in GDM pathogenesis. Similarly, we found elevated total NK cells and ILC1s **(Figure 2D, Supplementary Fig. 2A)** and increased frequencies of HLA-DR^+^ ILC1s **(Fig. 2G)** in peripheral blood of patients with a history of GDM and insulin treatment during pregnancy. Importantly, these immune cell alterations persisted over the 5-year follow-up period. Recent data highlighted the role of ILCs in modulating immune responses and influencing metabolic homeostasis (26). Particularly, adipose tissue ILC2s have been shown to promote metabolic homeostasis, adipose tissue “browning”, and systemic insulin sensitivity, protecting against obesity-induced metabolic disorders and type 2 diabetes (26). In line with these findings, we found significant associations between circulating frequencies of ILC2s and ISI, whereas total ILC1s and percent HLA-DR^+^ ILC1s were negatively correlated **(Fig. 3A-C)**. However, our work does not resolve the functional contribution of peripheral blood ILC2s in protecting against insulin resistance, and ILC subsets could not predict the risk of progression to prediabetes after GDM.

A limitation of our study is that we only report data from the first year postpartum (Visit 1) and the 5-year follow-up (Visit 3). Clinically apparent diabetes may take more than five years to manifest in women with a history of GDM, as previously suggested (43). However, future work will provide data from 10-year follow-up visits on the ongoing prospective PPSDiab study. An additional constraint is that we did not investigate GDM and healthy control patients during pregnancy, thus, precluding us from investigating pregnancy-related differences in ILCs. In contrast, data from the multicenter, prospective PREG study (NCT 04270578) suggest metabolic alterations and associations of pre-pregnancy BMI with IL-6 levels in late pregnancy in GDM patients as compared to healthy controls (44,45).

The precise mechanisms by which ILC populations contribute to the development of an impairment of glucose homeostasis remain to be fully elucidated. It is crucial to explore the interactions between ILCs and other immune cells, such as macrophages and T cells, as well as their impact on adipose tissue function and pancreatic beta-cell health. Understanding these relationships could provide deeper insights into the pathophysiology of GDM and its long-term consequences. We previously reported that frequencies of naïve and memory CD4^+^ and CD8^+^ T cells inversely correlated with measures for insulin sensitivity in humans. Moreover, percent naïve CD4^+^ and CD8^+^ T cells were significantly higher, whereas activated T cells and IL-6 levels were lower in insulin sensitive compared to insulin resistant obese individuals (46). The association between T cell senescence and insulin resistance was confirmed in both circulating and subcutaneous adipose tissue T cells of postmenopausal obese women (47). In the context of GDM, the activity and distribution of these cells may be altered, contributing to both the immediate metabolic disturbances observed during pregnancy and the long-term risk of glucose impairment. Key open questions remain regarding specific roles of both T cells and different ILC populations in the progression from GDM to type 2 diabetes, the crosstalk between immune and metabolic systems in obesity and insulin resistance, and the identification of biomarkers that could predict long-term glucose outcomes in women with a history of GDM. Future work should consider immune modulation as a potential target for therapeutic intervention in GDM and subsequent glucose impairment to pave the way for personalized approaches to preventing diabetes in this high-risk population. In summary, our study identifies sustained increase in circulating HLA-DR^+^ ILC1s in women with a history of GDM and insulin treatment during pregnancy as compared to GDM patients without insulin treatment and healthy controls, although these immune alterations were likely driven more by overweight/obesity and metabolic changes than by GDM itself. We demonstrate that activated ILC1s and NK cells correlate with measures of obesity and insulin resistance during the follow-up period. It will be of interest to identify human adipose ILC1 subsets in women with a history of GDM and determine whether these cells are dysregulated and can contribute to metabolic disorders. Future work will elucidate the specific roles of different ILC subsets in the progression from GDM to type 2 diabetes and focus on metabolic risk markers that could predict long-term glucose outcomes in women with a history of GDM.

## Abbreviations

GDM: Gestational diabetes mellitus
HOMA-IR: Homeostatic Model Assessment for Insulin Resistance
IADPSG: International Association of the Diabetes and Pregnancy Study Groups
ILCs: Innate lymphoid cells
ILC1s: Group 1 innate lymphoid cells
ILC2s: Group 2 innate lymphoid cells
ILCps: Innate lymphoid cell precursors
ISI: Insulin sensitivity index
OGTT: Oral glucose tolerance test
PBMC: Peripheral blood mononuclear cells
p.p.: postpartum
PPSDiab: Prediction, Prevention and Subclassification of Type 2 Diabetes

## Data availability statement

Data described in the manuscript will be available from the corresponding author upon reasonable request. All data supporting the findings of this study are available within the paper and its Supplementary Material.

## Ethics statement

This study was performed in line with the principles of the Declaration of Helsinki. Approval was granted by the Ethics Committee of the Ludwig-Maximilians-University (study ID 300-11). Written informed consent was obtained from all study participants.

## Author contributions

**Julia Sbierski-Kind:** Writing – original draft, Methodology, Conceptualization, Project administration, Data curation, Visualization, Formal analysis. **Stephan Schlickeiser:** Data curation, Visualization, Formal analysis. **Lorenzo Semeia:** Formal analysis. **Saori Harada:** Formal analysis. **Eleni Pappa:** Data curation. **Javier Villamizar Cujar:** Data curation. **Minh-Thuy Katschke:** Formal analysis. **Christina Gar:** Data Curation. **Andreas Lechner:** Supervision, Conceptualization, Funding acquisition. **Andreas Birkenfeld:** Supervision, Funding acquisition. **Uta Ferrari:** Conceptualization. Data curation. Formal analysis. **Jochen Seissler:** Conceptualization. Supervision. Funding acquisition.

## Acknowledgements

We thank all participants of the PPS-Diab study for their contribution to this project. We are also grateful to the diabetes care team of the Medizinische Klinik IV.

## Generative AI statement

The author(s) declare no Generative AI was used in the creation of this manuscript.

## Supplementary material

The Supplementary Material for this article can be found online at:

