## Supplementary material for "Association of overweight/obesity and insulin resistance with activation of circulating innate lymphoid cells in women after gestational diabetes mellitus"

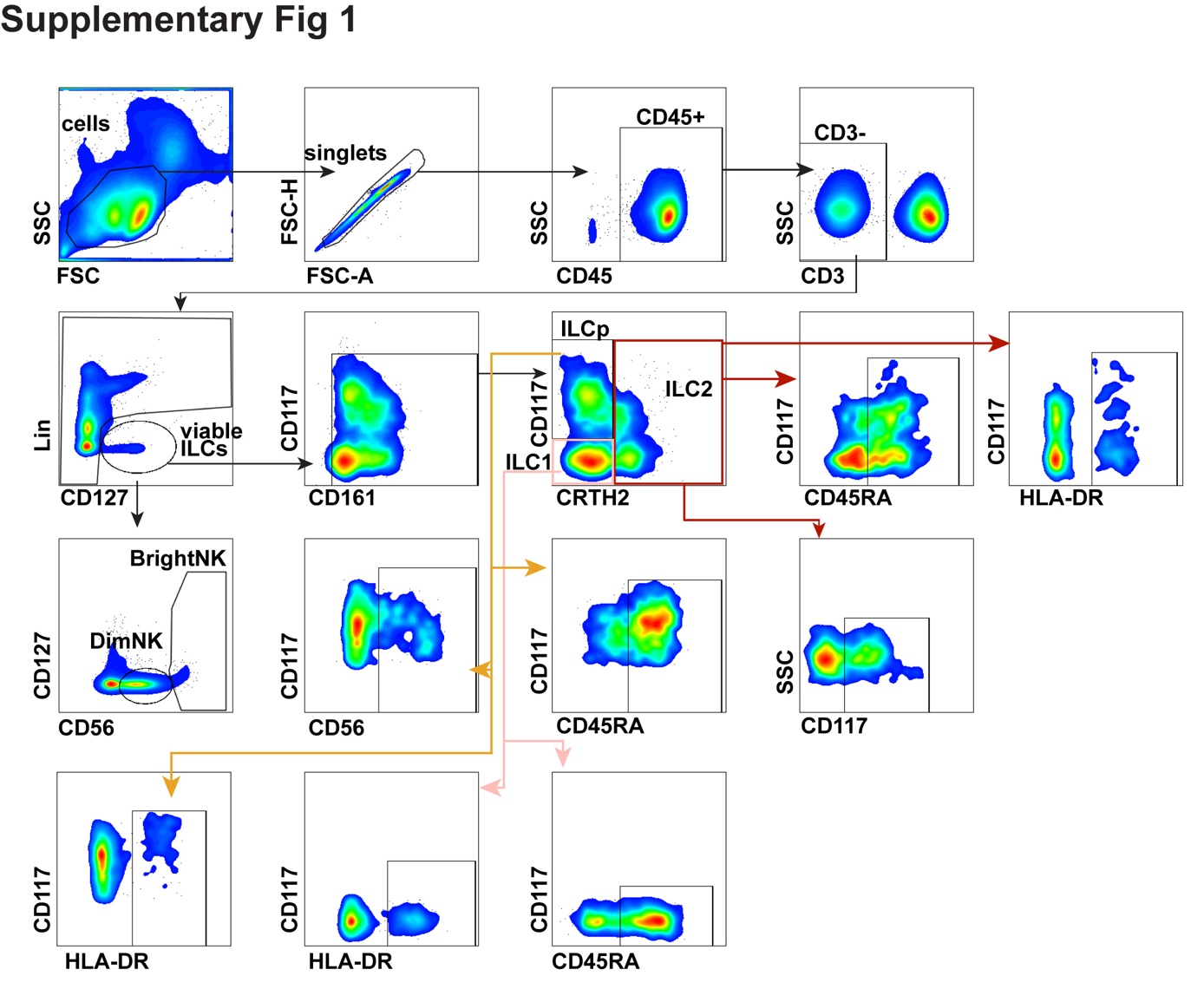

**Supplementary Figure 1 Flow cytometry gating scheme.** Representative flow cytometry gating scheme for analysis of innate lymphoid cells (ILCs) in peripheral blood mononuclear cells (PBMCs). Lineage markers contained CD1a, CD14, CD19, CD123, BDCA2, FceR1, CD34, CD94, TCRαβ, TCRγδ, FceR1, and dead cell marker.

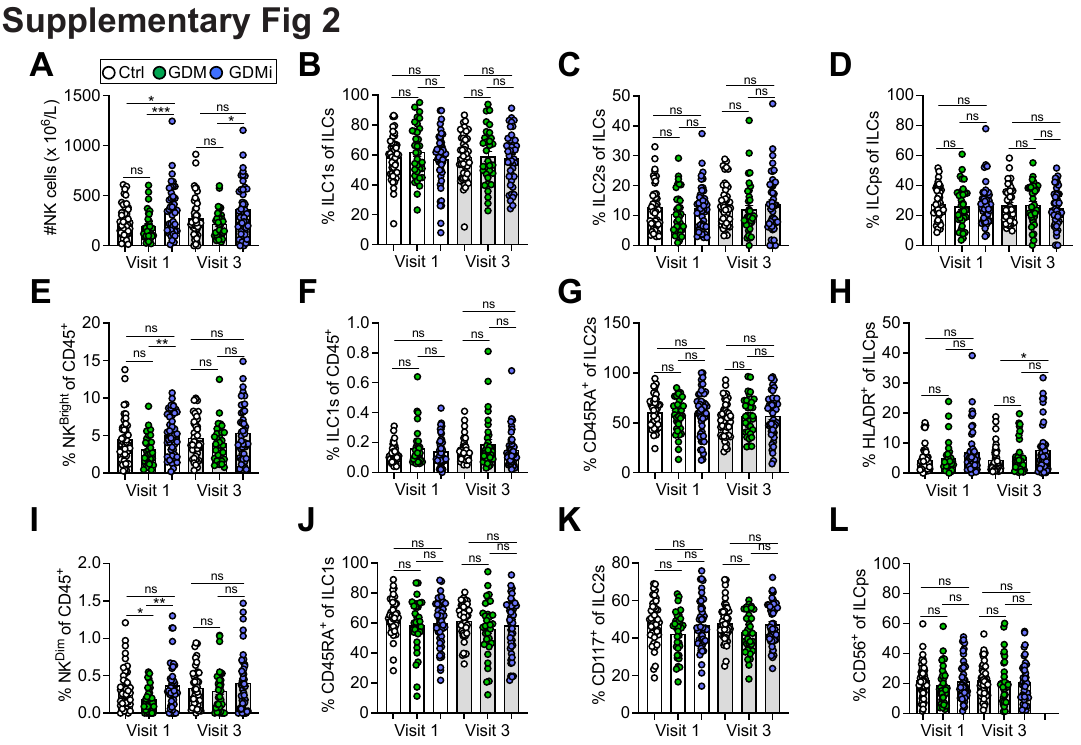

**Supplementary Figure 2 (A, E, I)** Quantifications (A) and percent (E, I) peripheral blood NK cells in healthy control females with normoglycemic pregnancies (Ctrl), females post gestational diabetes mellitus (GDM), and females post gestational diabetes mellitus with insulin treatment during pregnancy (GDMi) at Visit 1 (3-16 months postpartum) and at Visit 3 (58-66 months postpartum). **(B, F, J)** Percent peripheral blood group 1 innate lymphoid cells (ILC1) in Ctrl, GDM, and GDMi study participants at Visit 1 and Visit 3. **(C, G, K).** Percent peripheral blood group 2 innate lymphoid cells (ILC2) in Ctrl, GDM, and GDMi study participants at Visit 1 and Visit 3. **(D, H, L)** Percent peripheral blood innate lymphoid progenitors (ILCps) in Ctrl, GDM, and GDMi study participants at Visit 1 and Visit 3. Bar graphs indicate mean (±standard error), n=33-45 individuals per group, one-way ANOVA with Tukey post hoc test; *p ≤ 0.05, **p ≤ 0.01, ***p ≤ 0.001.

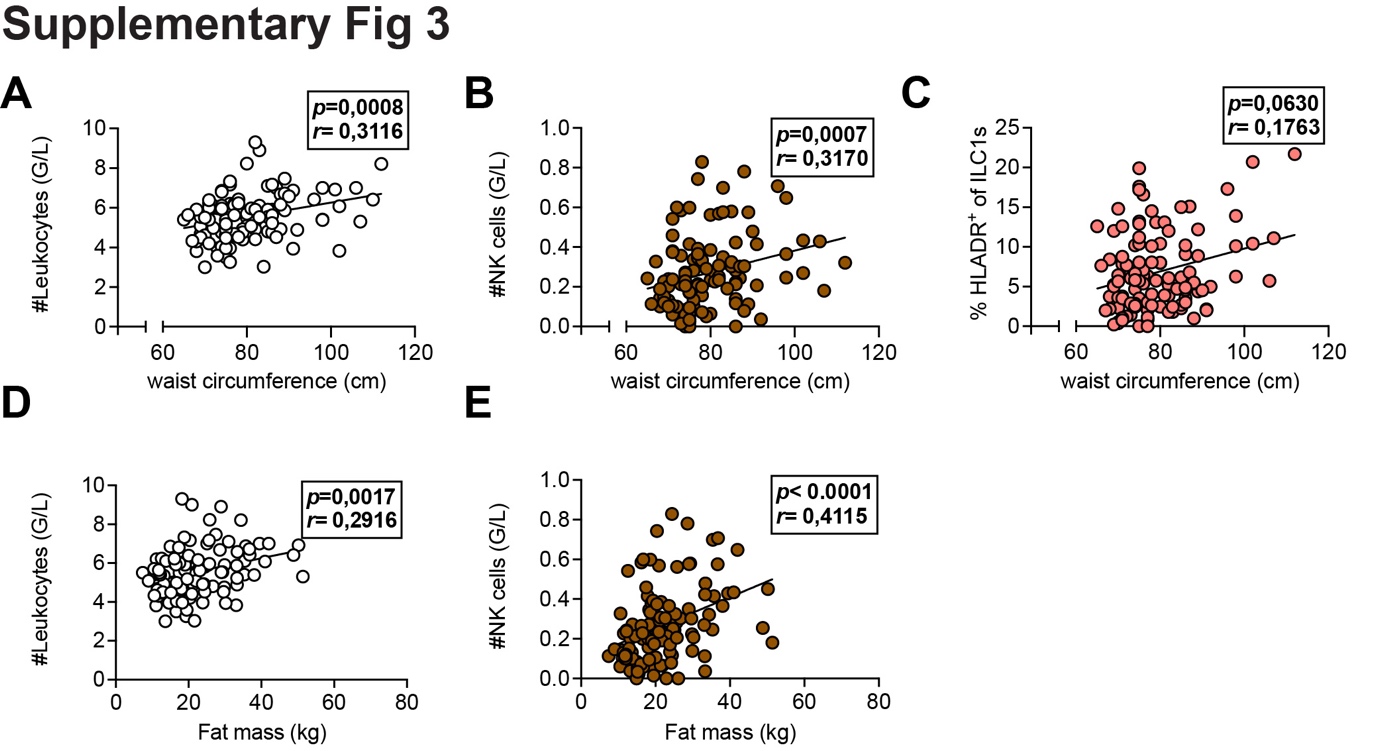

**Supplementary Figure 3** Associations of systemic leukocyte subpopulations with metabolic measures. Peripheral blood mononuclear cells (PBMCs) isolated from peripheral blood of healthy control females with normoglycemic pregnancies (Ctrl), females post gestational diabetes mellitus (GDM), and females post gestational diabetes mellitus with insulin treatment during pregnancy (GDMi) at Visit 3 (58-66 months postpartum). Subjects were analyzed with multiparameter flow cytometry and the resulting immune cell parameters were correlated with metabolic measures. **(A-C)** Correlations between leukocyte counts (A), total NK cells (B), and percent HLADR^+^ of ILC1s (C) and waist circumference. **(D, E)** Correlations between leukocytes (D) and total NK cells (E) with fat mass. n=33-45 individuals per group. Spearman correlation analyses were conducted.

**Supplementary Table 1** Follow-up characteristics of study participants at Visit 3.

**Supplementary Table 2** Antibody list for multiparameter flow cytometry.
