## Supplementary material for "Association of overweight/obesity and insulin resistance with activation of circulating innate lymphoid cells in women after gestational diabetes mellitus": Table S1

**Table 1** Follow-up characteristics of study participants at Visit 3

| Group | Control | GDM | GDMi | ***Adjusted p*-value (Ctrl. vs. GDM)** | ***Adjusted p*-value (Ctrl. vs. GDMi)** | ***Adjusted p*-value (GDM vs. GDMi)** |
|---|---|---|---|---|---|---|
| No. of subjects | 45 | 33 | 42 | | | |
| Age (years) | 39.62 ± 4.17 | 39.42 ± 3.99 | 40.52 ± 4.21 | 0.833 | 0.319 | 0.254 |
| BMI ($kg/m^2$) | 23.92 ± 4.24 | 22.93 ± 3.47 | 26.38 ± 5.77 | 0.277 | 0.026 | 0.003 |
| Waist circumference (cm) | 78.4 ± 8.78 | 77.24 ± 7.59 | 83.98 ±11.32 | 0.544 | 0.012 | 0.004 |
| Fat mass (kg) | 21.63 ± 8.29 | 18.74 ± 6.79 | 25.72 ± 9.98 | 0.105 | 0.041 | 0.001 |
| HbA1c (%) | 5.18± 0.29 | 5.33 ± 0.34 | 5.41 ± 0.31 | 0.035 | 0.000 | 0.321 |
| HDL cholesterol (mg/dl) | 67.58 ± 13.74 | 67.30 ± 15.38 | 62.48 ± 13.69 | 0.934 | 0.086 | 0.155 |
| LDL cholesterol (mg/dl) | 99.24 ± 28.06 | 95.45 ± 25.45 | 102.50± 29.62 | 0.541 | 0.599 | 0.280 |
| Triglycerides (mg/dl) | 81.22 ± 31.67 | 70.82 ± 33.62 | 103.17 ± 56.20 | 0.166 | 0.026 | 0.004 |
| TSH (uIU/ml) | 1.93 ± 0.98 | 1.65 ± 0.75 | 1.86 ± 0.99 | 0.176 | 0.756 | 0.309 |
| Leukocytes ($10^9$/L) | 5.38 ± 1.12 | 5.27 ± 1.13 | 6.23 ± 1.51 | 0.664 | 0.003 | 0.003 |
| Glucose metabolism at Visit 3 (%) | | | | | | |
| NGT | 37 (82.2) | 23 (69.7) | 23 (57.2) | | | |
| IFG | 7 (15.6) | 5 (15.1) | 9 (21.4) | | | |
| IGT | 1 (2.2) | 3 (9.1) | 4 (9.5) | | | |
| IFG+IGT | 0 (0) | 2 (6.1) | 5 (11.9) | | | |
| T2DM | 0 (0) | 0 (0) | 0 (0) | | | |

Data are presented as means (± standard deviation). Adjusted p-values were obtained from post-hoc tests adjusted for multiple comparisons following a one-way analysis of variance (ANOVA) or Kruskal-Wallis test. Visit 3 (58-66 months postpartum). GDM, gestational diabetes mellitus. BMI, body mass index. IFG, impaired fasting glucose. IGT, impaired glucose tolerance. NGT, normal glucose tolerance.
