## Supplementary material for "Association of overweight/obesity and insulin resistance with activation of circulating innate lymphoid cells in women after gestational diabetes mellitus": Table S2

**Table S1.** Antibodies for flow cytometry.

| REAGENT or RESOURCE | SOURCE | IDENTIFIER |
|---|---|---|
| **Antibodies** | | |
| Anti human CD1a FITC (clone HI149) | Biolegend | Cat# 300103 |
| Anti human CD14 FITC (clone TuK4) | Life Technologies | Cat# MHCD14014 |
| Anti human CD19 FITC (clone 4G7) | BD Biosciences | Cat# 345776 |
| Anti human CD123 FITC (clone 6H6) | Biolegend | Cat# 306013 |
| Anti human BDCA2 FITC (clone AC144) | Miltenyi | Cat# 130-090-510 |
| Anti-human FceER1a FITC (clone AER-37) | Biolegend | Cat# 334607 |
| Anti-human CD34 FITC (clone 581) | Biolegend | Cat# 343503 |
| Anti-human CD94 FITC (clone DX22) | Biolegend | Cat# 305504 |
| Anti-human TCRab FITC (clone IP26) | Biolegend | Cat# 306705 |
| Anti-human TCRgd FITC (clone B1) | Biolegend | Cat# 331207 |
| Live/Dead Green FITC | Life Technologies | Cat# L23101 |
| Anti-human CD45RA APC (clone HI100) | Biolegend | Cat# 304111 |
| Anti-human CD161 BV 605 (clone HP-3G10) | Biolegend | Cat# 339915 |
| Anti-human CD45 BV 650 (clone HI30) | Biolegend | Cat# 304043 |
| Anti-human CD56 BV 711 (clone HCD56) | Biolegend | Cat# 318335 |
| Anti-human CD3 BV 785 (clone OKT3) | Biolegend | Cat# 317329 |
| Anti-human CRTH2 PE-CF954 (clone BM16) | BD Biosciences | Cat# 563501 |
| Anti-human CD117 PE-Cy5.5 (clone 104D2D1) | Beckman Coulter | Cat# A66333 |
| Anti-human CD127 PE-Cy7 (clone R34.34) | Beckman Coulter | Cat# A64618 |
| Anti-human SLAMF1 BV421 (clone A12) | BD Biosciences | Cat# 562875 |
| Anti-human HLADR BV510 (clone L243) | Biolegend | Cat# 307646 |
